# The iDiabetes Platform: Enhanced Phenotyping of Patients with Diabetes for Precision Diagnosis, Prognosis and Treatment - study protocol for a cluster-randomised controlled study in Tayside, Scotland

**DOI:** 10.1101/2024.03.19.24304468

**Authors:** YeunYi Lin, Damien Leith, Michael Abbott, Rachael Barrett, Samira Bell, Tim Croudace, Scott G Cunningham, John F Dillon, Peter T Donnan, Albert Farre, Rodolfo Hernández, Chim Lang, Stephanie McKenzie, Ify R Mordi, Susan Morrow, H Cameron Munro, Mandy Ryan, Deborah J Wake, H Wang, Mya Win, Ewan R Pearson, the iDiabetes Study Team

## Abstract

**Introduction and Aim:** Diabetes is a global health emergency with increasing prevalence and diabetes-associated morbidity and mortality. One of the challenges in optimising diabetes care is translating research advances in this heterogenous disease into clinical care. A potential solution is the introduction of precision medicine approaches into diabetes care.

We aim to develop a digital platform called ‘intelligent Diabetes’ (iDiabetes) to support a precision diabetes care model in Scotland and assess its impact on the primary composite outcome of all-cause mortality, hospitalisation rate, renal function decline and glycaemic control.

**Methods and Analysis:** The impact of iDiabetes will be evaluated through a cluster-randomised controlled study, recruiting up to 22,500 patients with diabetes. Primary care general practices (GP) in the National Health Service Scotland Tayside Health Board are the units (clusters) of randomisation. Each primary care GP will form one cluster (approximately 400 patients per cluster), with up to 60 clusters recruited. Randomisation will be to *iDiabetes (guideline support)*, *iDiabetesPlus* or *usual diabetes care* (control arm). Patients of participating primary care GPs are automatically enrolled to the study when they attend for their annual diabetes screening or are newly diagnosed with diabetes. A composite hierarchical primary outcome, evaluated using Win-Ratio statistical methodology, will consists of (I) all-cause mortality, (II) all-cause hospitalisation rate, (III) proportion with >40% eGFR reduction from baseline or new development of end-stage renal disease, (IV) proportion with absolute HbA1C reduction >0.5%. Outcomes will be evaluated after a 2-year median follow-up period. Comprehensive qualitative and health economic analyses will be conducted, assessing the cost-effectiveness, budget impact and user acceptability of the iDiabetes platform.

**Ethics and Dissemination:** This study was reviewed by the NHS HRA and approved by East of Scotland Research Ethics Committee (reference:23/ES/0008). Study findings will be disseminated via publications, presented at scientific conferences and shared with patients and the public on the study website and social media.

**Study registration** ISRCTN18000901

**Study Sponsor** University of Dundee, no. 2-026-22. Contact

**Protocol version:** V3.0, 22/09/2023

**ARTICLE SUMMARY:** Strengths

- iDiabetes is a novel precision medicine platform which is the first of its kind to evaluate a precision diabetes care model in a controlled trial
- Real-world application in an existing healthcare system, with the entire regional diabetes population eligible for enrolment – results are therefore likely to be generalisable and the approach could be scaled up to a national level
- The study analysis utilises a mixed-method approach allowing a comprehensive evaluation of all aspects of the precision medicine platform including impact on clinical outcomes, usability for both patients and clinicians, cost-effectiveness analysis, budget impact analysis and patient preferences.

**Limitations:**

- Study enrolment takes place when patients attend their routine clinical diabetes review - consequently a subpopulation of patients with limited healthcare access may be excluded
- The patient-facing platform is only available in an English version and can only be accessed digitally, therefore patients with limited English or digital illiteracy may not benefit from the intervention to its full potential

## INTRODUCTION

### Background

Diabetes is increasing in prevalence worldwide, giving rise to a spectrum of associated co-morbidities including cardiovascular, renal and liver complications and posing a significant global health crisis. These complications, along with the burden of living with a chronic illness, contribute to impaired quality of life, disability and ultimately increased mortality in people living with diabetes. (1) Public health strategies have been unsuccessful at controlling the epidemic and current pharmacological therapy is often sub-optimal – focussed largely on glycaemic regulation, with prescribing practice sometimes influenced by non-medical factors such as drug cost. As a result, opportunities to target therapy choice to underlying biological mechanisms of diabetes and relevant comorbidities may be overlooked. (2)

Recognising the highly heterogenous nature of diabetes, the concept of precision diabetes medicine has garnered attention in modern diabetes care. Research breakthroughs in diabetes in recent decades creates the potential for a revolution in care, incorporating a more comprehensive analysis of multiple biological factors, including genomic variations and metabolic pathways, with environmental risk factors for individual patients. (2) For example, recent research has shed light on the heterogeneity of type 2 diabetes (T2D) which has traditionally been viewed as a uniform pathology. This heterogeneity exists as a continuum and is associated with both phenotypic and genotypic variations, resulting in differential disease progression, end-organ complications and treatment response. (3, 4) Additionally, predictive algorithms derived from patient level characteristics within a UK-based population database demonstrated that this phenotypic heterogeneity could be used to guide treatment choices (e.g. for choosing between SGLT2 inhibitor *vs* DPP4 inhibitor) to optimise glycaemic control.(5) More recently, clinical diagnostic models incorporating clinical features and biomarkers (such as pancreatic autoantibodies and Type 1 Diabetes Genetic Risk Score) may allow enhanced differentiation between type 1 diabetes (T1D) and T2D at the time of diagnosis. (6) Similarly, the identification of monogenic forms of diabetes has allowed for cessation of unnecessary diabetes treatment in patients with *GCK*-MODY and spared patients with other forms, such as neonatal diabetes and *HNF1A*-MODY, from unnecessary insulin injections. (7–9) However, incorporating these advances into routine clinical care, translating them to direct benefit to patients, is challenging and can be slow. For example, despite being well described in the literature, it has been estimated that approximately 80% of monogenic diabetes cases remain undiagnosed, highlighting the need for more effective ways to translate research outcomes into routine clinical care.(10)

Introducing precision medicine in diabetes care, as outlined in the consensus American Diabetes Association (ADA)/European Association for the Study of Diabetes (EASD) Precision Diabetes Medicine Initiative (PDMI) report, requires the transformation of the current care approaches.(11) Using a range of data, a more refined diabetes care model should be developed which encompasses all aspects of the disease – disease prevention, timely diagnosis, personalised treatment, individualised prognosis and appropriate monitoring of diabetes.

### Implementing precision diabetes care in Scotland

We aim to develop and assess an innovative “intelligent Diabetes” (iDiabetes) platform to support and implement a precision diabetes care model, delivered at scale, to patients with diabetes managed in both primary and secondary care in the Tayside Health Board region of Scotland. The platform aims to facilitate precision diagnosis and promote the selection of the optimal treatments by healthcare professionals, while ensuring that these diagnoses and treatment recommendations are effectively communicated to patients.

This study will compare two different arms of the iDiabetes platform to usual diabetes care. The *iDiabetes (guideline support)* arm will utilise routinely collected clinical and biochemical data to produce individualised treatment recommendations.

The *iDiabetesPlus* arm will combine routinely collected clinical and biochemical data with additional biochemical and genotypic testing to determine a patient’s risk of current and future end-organ diabetes complications and predicted glycaemic response to different pharmacotherapies. This information will be used to generate enhanced individualised treatment recommendations.

The treatment recommendations generated in both arms of the iDiabetes platform will allow healthcare professionals to make better-informed, individualised evidenced-based treatment decisions, thereby improving management for each patient. If demonstrated to improve patient care and to be cost-effective, the vision is for iDiabetes to be the standard of care for patients with diabetes across Scotland.

### Study objectives

#### Primary objective

- Develop a precision medicine platform for diabetes care and assess its impact on the primary composite outcome (mortality rate, hospitalisation rate, decline in renal function and glycaemic control) of patients with diabetes

#### Secondary objectives

- Assess the impact of a precision medicine platform on the individual components of the composite outcomes of patients with diabetes
- Evaluate the impact of a precision medicine platform on hospitalisation rate secondary to heart failure
- Evaluate patients’ adherence to antidiabetic medications, cholesterol-lowering medications and antihypertensives
- Evaluate the impact of iDiabetes platform treatment recommendations on the rate of severe hypoglycaemia
- Evaluate whether recommendations from iDiabetes platform improve healthcare professionals’ compliance with guideline recommendations during prescription of antidiabetic medications

Additional exploratory objectives will include further assessment of the impact of iDiabetes on end-organ diabetes-associated complications, qualitative and costeffectiveness analysis of the platform. A full list of exploratory objectives can be viewed in the Supplemental File.

## METHODS

### Study design

This is a three-arm cluster-randomised controlled study (two iDiabetes arms and usual diabetes care control arm), with a nested qualitative study and economic evaluation. The inclusion of two different arms within the iDiabetes platform allows evaluation of the clinical utility and cost-effectiveness of the addition of guideline-based recommendations *(iDiabetes (guideline support))* to patients’ usual diabetes care as well as the additional value gained from precision diagnosis and treatment resulting from the addition of enhanced phenotyping and genotyping to guideline support *(iDiabetesPlus arm)*.

### Randomisation

Each primary care general practice (GP) within the National Health Service (NHS) Tayside Scottish Health Board region will form one of up to 60 clusters. NHS Tayside provides primary healthcare to approximately 417,650 people in the northeast of Scotland, in both urban and rural communities within the region.(12) Practices will be stratified by population size (<6000 or ≥6000 registered patients) and index of deprivation deciles (1-3 high deprivation, 4-7 medium, 8-10 low).(13) Clusters will be randomised to one of the following three arms: *iDiabetes (guideline support)*, *iDiabetesPlus* or *usual diabetes care* [see Figure 1]. All 60 clusters (approximately 400 patients per cluster) will undergo initial randomisation to the iDiabetes platform or *usual diabetes care* arms in a 2:1 ratio. Clusters initially allocated to the iDiabetes platform will undergo a second randomisation to *iDiabetes (guideline support)* or *iDiabetesPlus* in a 1:1 ratio, resulting in overall cluster allocation of 1:1:1 for *iDiabetes (guideline support)*, *iDiabetesPlus* and *usual diabetes care* arms.

**Figure 1:**
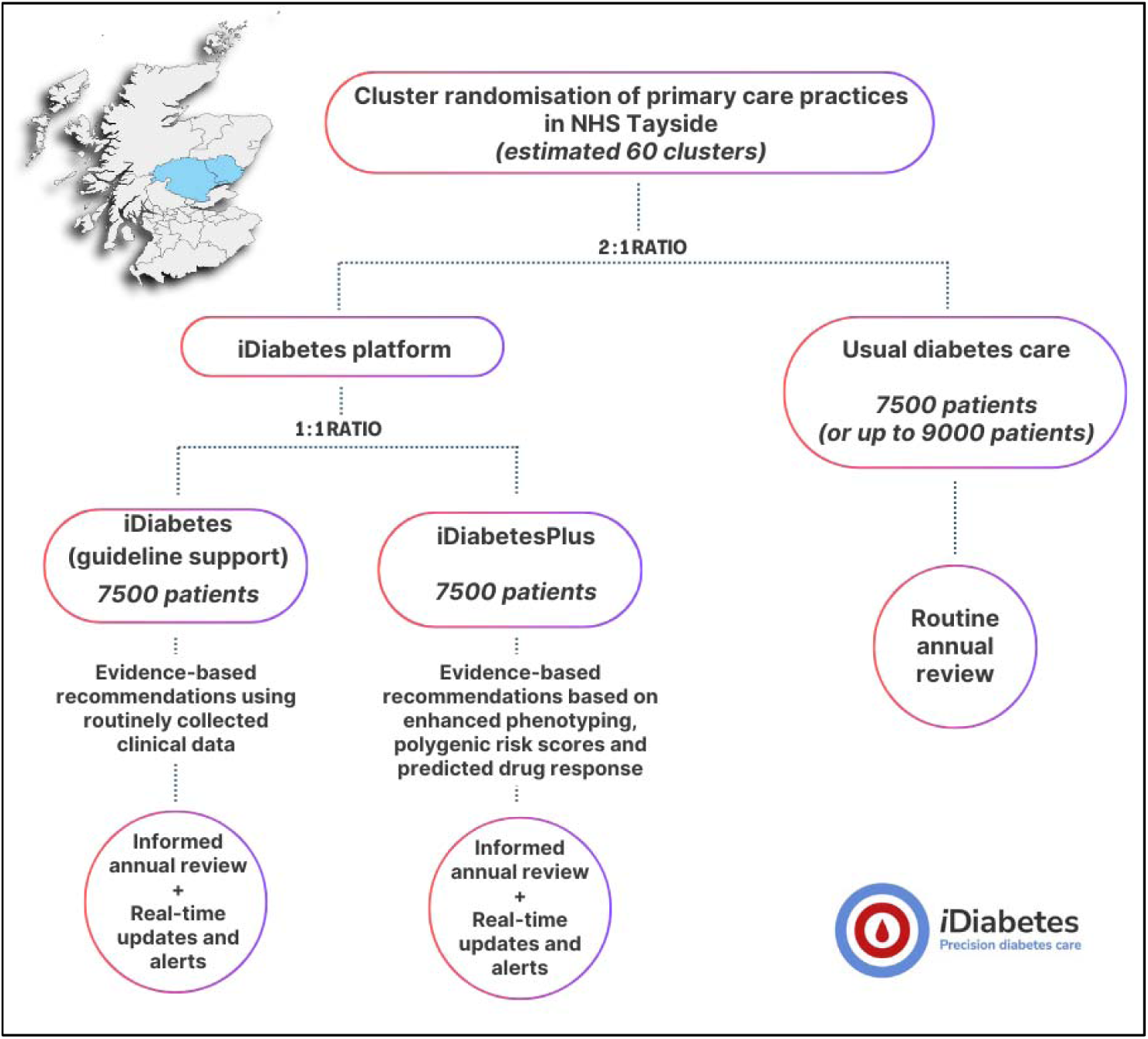
Cluster randomisation with each GP practice forming a cluster. Map at the top left corner of the figure shows the geographical area covered by NHS Tayside health board in Scotland (generated via paintmaps.com).

The randomisation will be created by the UKCRC registered Tayside Clinical Trials Unit (TCTU). The study participants, primary care GPs and study clinicians will not be blinded. Blinding would not be possible as test results will be accessible to healthcare professionals and patients as part of the iDiabetes platform design, and the tests undertaken will be determined by the study arm therefore this will make the study arm known. Additionally, specific consent is required for genetic testing which is only performed in the *iDiabetesPlus* arm. There are no planned criteria for stopping or switching a patients’ enrolment in an iDiabetes intervention arm.

### Recruitment of primary care GPs

All primary care GPs in NHS Tayside will be invited to participate in the study. The study team will actively promote the study and engage with primary care GPs to encourage their participation. This will ensure that the adequate sample size is met at cluster level.

### Recruitment of participants and follow-up period

Patients will be enrolled into the study when they attend for their routine annual diabetes screening appointment or are newly diagnosed with diabetes in primary care during the study period. Recruitment will stop when 7,500 patients, with diabetes of any type, have been enrolled in each arm. Recruitment into the *usual diabetes care* arm will be allowed to continue up to a maximum of 9000 patients if this arm reaches completion earlier than the *iDiabetes (guideline support)* or *iDiabetesPlus* arms. It is anticipated that the total recruitment period will be 12 – 15 months.

The planned study follow-up period will be until a median of 2 years of follow up has been completed across all patients recruited into the study.

It is anticipated that study recruitment will begin in August 2024.

### Eligibility criteria

#### Inclusion criteria

- Patient registered with an NHS Tayside Health Board primary care GP practice which has signed up and agreed to commit for the duration of the study
- Diagnosis of any diabetes type
- Aged ≥18 years at time of diabetes annual review during the study period

#### Exclusion criteria

There will be no exclusion criteria. All patients identified as meeting the inclusion criteria will be included in the study.

### Intervention

The iDiabetes platform will form the central intervention in this study. The iDiabetes platform will incorporate patient data and facilitate the generation of recommendations based on the most recent evidence for each patient. The recommendations will be accessible by both healthcare professionals, via the iDiabetes dashboard, and by patients themselves, via My Diabetes My Way (MDMW). MDMW is an interactive online patient portal used by patients with diabetes in Scotland. It provides them with information about their condition and secure access to their own diabetes-related healthcare records, including test results.(14) For all patients enrolled in the study there will be no restriction or implication for any other concomitant care or interventions provided by the patients’ usual care team as the iDiabetes platform is a tool to support decision making only.

#### iDiabetes (guideline support)

Patients will attend their annual diabetes review and be assessed as per usual care including routine diabetes blood testing [see Table 1]. The iDiabetes platform will generate automated, individualised treatment recommendations, using routine clinical data and blood test results from annual appointments. Recommendations will be determined by the latest treatment guidelines such as *American Diabetes Associations’ Standard of Care for Diabetes* and *National Institute for Health and Care Excellence (NICE) Clinical Knowledge Summaries.*(15, 16) Recommendations on optimisation of medications for associated co-morbidities will also be provided where a patient has renal or cardiovascular disease. For example, advice will be given to optimise ACE inhibitor or angiotensin receptor blocker for patients with proteinuria or optimise statins for patients with cardiovascular.

**Table 1:**
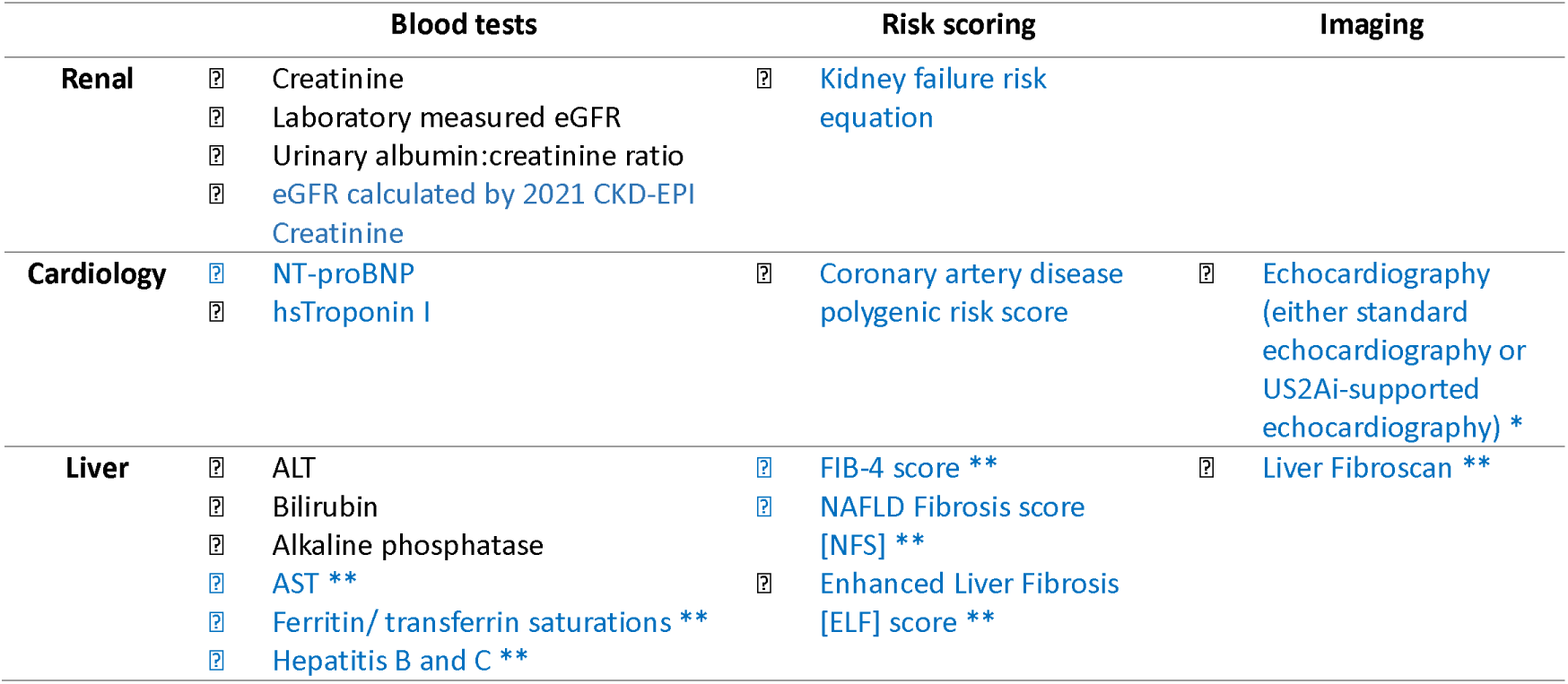
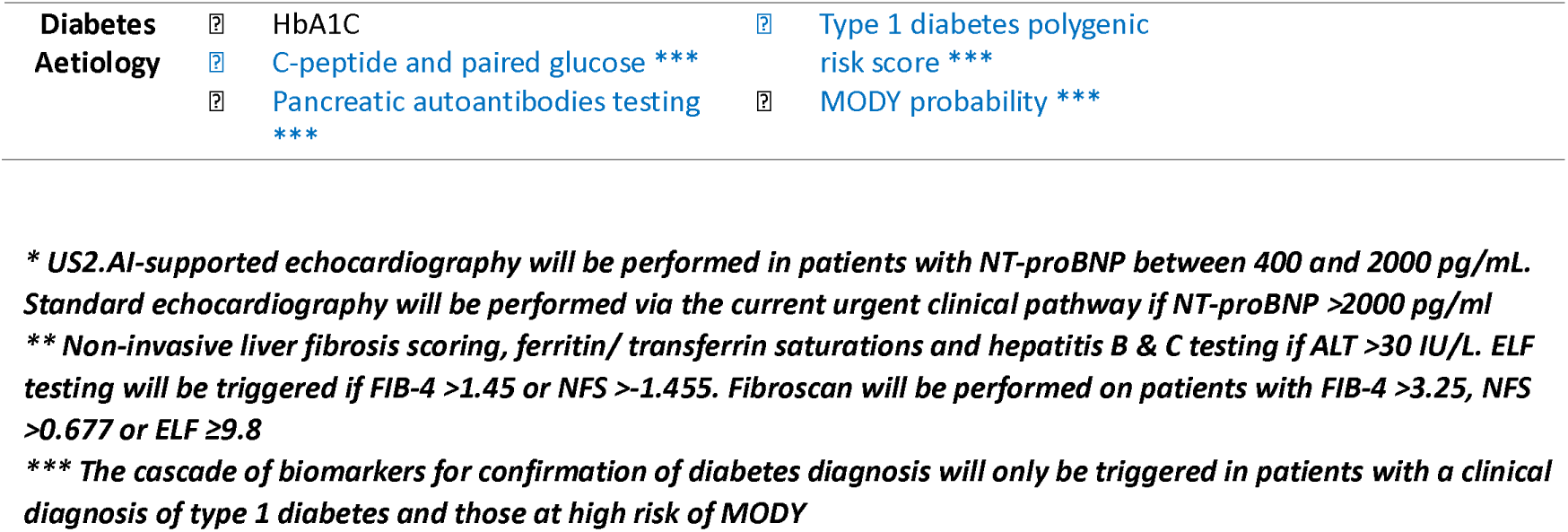
Blood tests, risk scoring systems and imaging tests performed by iDiabetes platform. Tests in blue will be performed in *iDiabetesPlus* arm only.

#### iDiabetesPlus

Patients will undergo routine assessments at their annual diabetes review in the same way as *iDiabetes (guideline support)*. However, in addition to the routine diabetes blood testing, *iDiabetesPlus* will incorporate additional reflexive laboratory testing including C-peptide concentration, cardiac risk biomarkers +/- echocardiography, non-invasive liver fibrosis scoring +/- Fibroscan and individual genotyping (to allow for cardiovascular and type 1 diabetes genetic risk scoring) [see Table 1]. Enhanced phenotyping and genotyping will allow for improvement in diagnostic accuracy of diabetes type and determination of current and future risk of end-organ complications. Recommendations on optimisation of medications for associated co-morbidities will be provided in the same way as *iDiabetes (guideline support)* with additional recommendations for patients with liver disease. Further to this, risks of myocardial infarction and all-cause mortality will be predicted for patients with T2D using models developed by MyWay Digital Health (MWDH). Treatment recommendations for each patient will be evidence-based and determined according to their competing cardiorenal or liver risk, risk of hypoglycaemia and predicted diabetes drug treatment response using a treatment selection algorithm. All recommendations will be accessible by the patient and their diabetes care team [see Figure 2].

**Figure 2:**
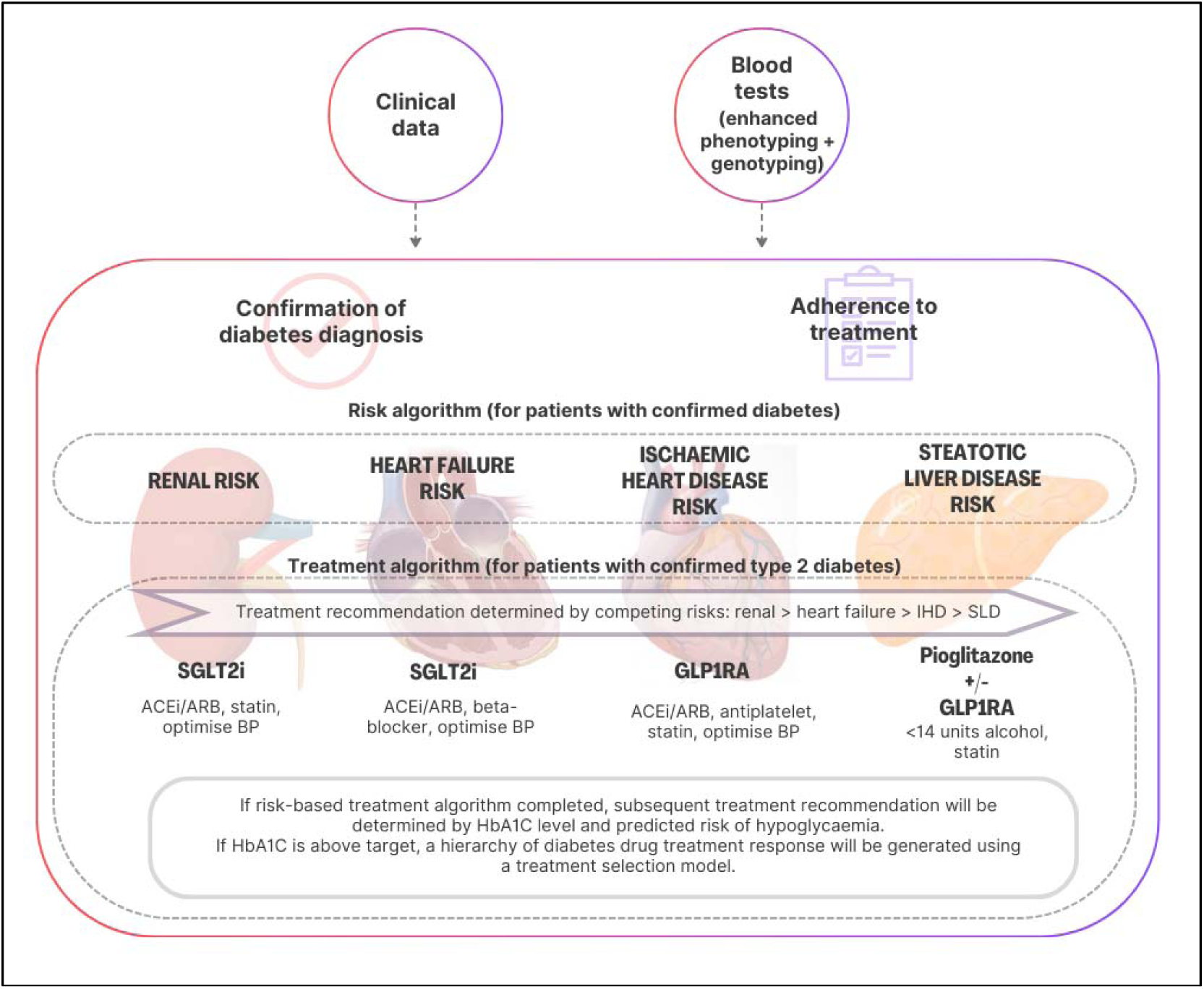
An overview of iDiabetesPlus. ACEi: angiotensin-converting enzyme inhibitor; ARB: angiotensin receptor blocker; BP: blood pressure; IHD: ischaemic heart disease; GLP1RA: glucagon-like-peptide 1 receptor agonist; SGLT2i: sodium-glucose transporter 2 inhibitor; SLD: steatotic liver disease; 1 unit of alcohol = 8g of ethanol.

#### Usual diabetes care

This will form a control arm. Patients will attend their usual annual diabetes appointment and be managed as per the usual care by their diabetes care provider, with no additional intervention from the iDiabetes platform or study team.

### Development of iDiabetes platform

The iDiabetes platform will be made possible through the development of a novel software called iDiabetes IQ engine which will be registered as a Class 1 medical device under the current UK Medical Device Directive according to ISO standards [see Figure 3].(17) Comprehensive information regarding data source and exchange for the software will be provided in a separate article on the programming of iDiabetes IQ engine. In this section, we provide a brief overview of the steps involved in developing the software.

**Figure 3:**
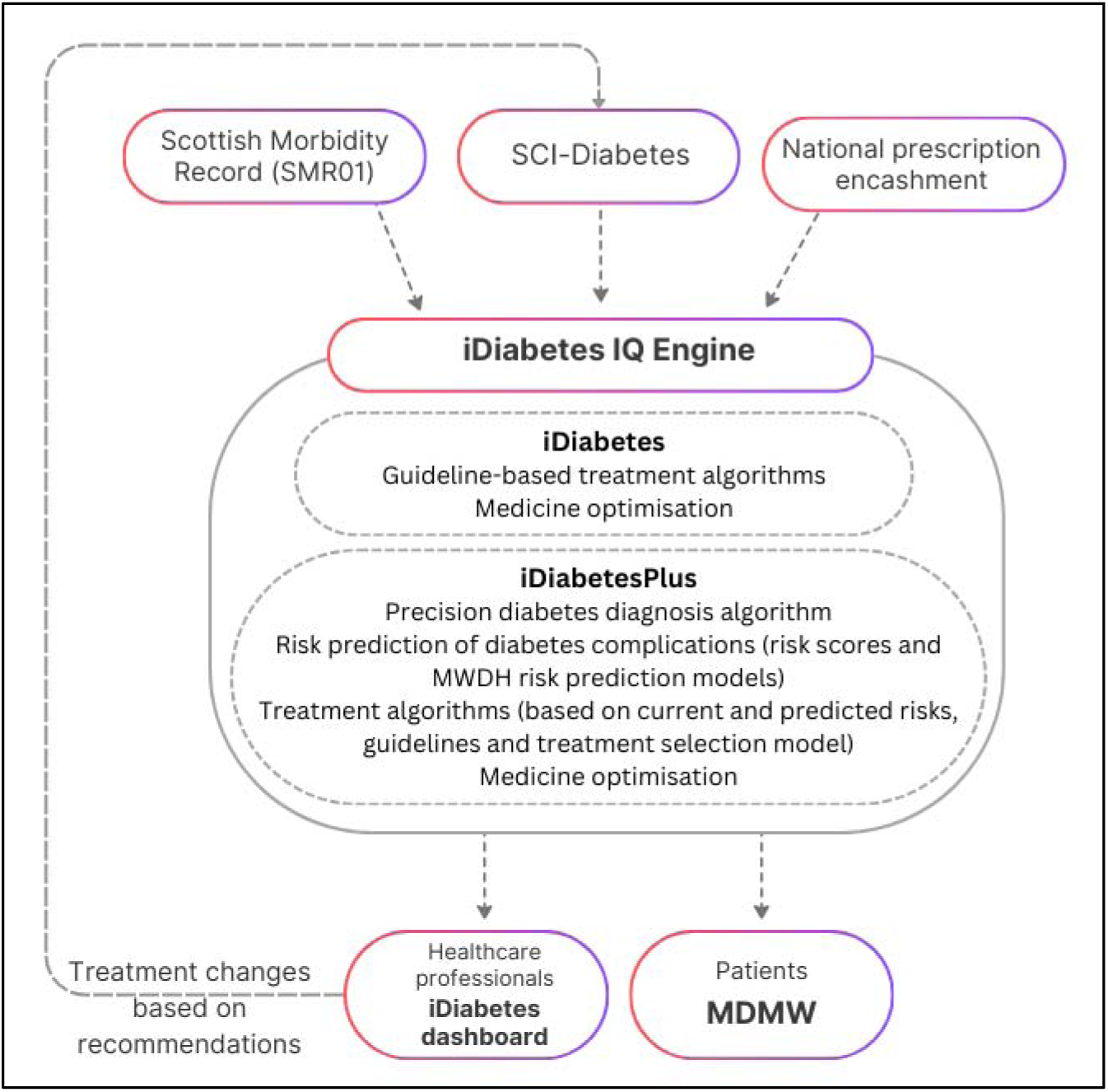
An overview of the iDiabetes IQ engine. MWDH: *MyWay Digital Health*; MDMW: *My Diabetes My Way*

#### Development of algorithms

The iDiabetes IQ engine software will consist of a series of rules-based algorithms encompassing three main themes: diagnosis of diabetes, risk stratification of end-organ complications and diabetes treatment recommendations. The *iDiabetes (guideline support)* and *iDiabetesPlus* algorithms were developed from a collaboration among expert clinicians utilising the latest guidelines, evidence and expert opinion.

#### From algorithms to iDiabetes dashboard content

The algorithm pathways were first converted into ‘pseudocode’ (a readable, but detailed description of what the software should do) and the ‘pseudocode’ translated into digital code by the iDiabetes software developers. iDiabetes dashboard contents were created which included explanatory text and links to external patient and clinician resources. A dashboard explanatory text was created for every recommendation generated by the algorithm. These will be viewable by healthcare professionals during clinic reviews. A lay version of the content was developed for patients to access via My Diabetes My Way.

#### iDiabetes IQ engine performance testing

To ensure that the iDiabetes IQ engine is functional and reliable, user acceptance testing was conducted to test its performance. A large ground truth dataset was created, with simulated patient data created to match every possible recommendation output. “Ground truth” patients were run through the algorithm to ensure all outputs matched the expected algorithm output. A subsequent full end-to-end testing, using an anonymised real-world dataset extracted from SCI-Diabetes, was performed. Any disparities between expected and observed outputs was fully scrutinised and where necessary the software code updated. A pilot phase on the roll out of the iDiabetes platform will facilitate a final period of real-world platform testing prior to the commencement of the full study.

#### Data input

Data input to the iDiabetes IQ engine will be sourced from SCI-Diabetes, an integrated clinical system with linked electronic health records from primary and secondary care, specialist screening services and laboratory systems for all patients with diabetes across NHS Scotland.(18) All patients’ clinical data and blood test results, both historical and that obtained during recruitment (when attending annual diabetes clinic appointment) will be available within the SCI-Diabetes system.

### Study outcomes

#### Primary outcome

A composite hierarchical outcome (in decreasing order of clinical importance):

1. All-cause mortality
2. All-cause hospitalisation rate
3. Proportion with >40% eGFR reduction from baseline, or new development of end-stage renal disease (ESRD)
4. Proportion with absolute HbA1C reduction >0.5% (>5.5mmol/mol)

#### Secondary outcomes

1. All-cause mortality rate
2. All-cause hospitalisation rate
3. Proportion with >40% eGFR reduction from baseline, or new ESKD
4. Proportion with absolute HbA1C reduction >0.5% (>5.5mmol/mol)
5. Hospitalisation rate secondary to heart failure
6. Drug adherence rate (medication possession ratio)
7. Rate of severe hypoglycaemia
8. Proportion of patients treated according to guidelines

Outcomes which can be measured in the *usual diabetes care* arm as well as the iDiabetes intervention arms have been selected to allow comparison. Therefore, in decreasing order of importance, we rank: 1) all-cause mortality, as failure to consider mortality can introduce issues of competing risk; 2) all-cause hospitalisations, as these are common and a major burden to patients and healthcare systems; 3) renal function decline, as this is a major morbidity in diabetes - a fall in eGFR of >40% is in keeping with clinical trials focusing on renal outcomes as the endpoint; 4) HbA1c reduction >0.5% (5.5mmol/mol), as HbA1c is a major driver of both micro and macrovascular disease in diabetes. HbA1c is not ranked higher despite its importance in diabetes as many treatment recommendations provided by iDiabetes (e.g. for HF, CAD, Renal disease, SLD) are HbA1c independent, in which HbA1c lowering is not our primary goal. All items in the composite will also be analysed and reported individually as secondary outcomes. All outcomes will be measured at individual patient level.

Additional exploratory outcomes will include further assessment of the impact of iDiabetes on end-organ diabetes-associated morbidity and diabetes treatment and a full qualitative and cost-effectiveness analysis of the platform. A full list of exploratory outcomes can be viewed in the Supplemental File.

## DATA COLLECTION, MANAGEMENT AND ANALYSIS

### Study Assessments

There will be no research specific study visits for primary and secondary outcomes as data will be collected from routine assessments during annual diabetes review and patient consent will be undertaken at a cluster level (see *iDiabetes Study Consent* below).

### Quantitative data collection and management

Blood and urine specimens will be taken from all patients during annual diabetes review appointments as part of routine clinical care. Additional testing will be performed on specimens from *iDiabetesPlus* clusters through a reflexive laboratory cascade system if criteria are met (see Table 1). Data will be collected for study outcome evaluation through routine data linkage, with clinical records and prescription encashment records used to evaluate medications adherence. All data will be analysed within a digital Trusted Research Environment (TRE) within the NHS Tayside & Fife Health Informatics Centre. All patient identifiable information will be anonymised within the TRE to ensure patient confidentiality. Data from patients in the study will be linked to their electronic medical records for long-term follow-up for up to 15 years.

The data management plan for the study was developed according to the Tayside Medical Science Centre standard operating procedure for data management.(19) As the iDiabetes platform is designed to integrate with routine diabetes clinical care, no efforts will be made to promote patient retention or completion of follow-up annual review appointments who would not ordinarily attend, to reflect real life circumstances.

### Qualitative data collection and management

This nested qualitative study will explore the views and experiences of those delivering and receiving iDiabetes and iDiabetesPlus care, using semi-structured interviews with patients and health professionals involved in the implementation and use of the iDiabetes platform and the delivery of iDiabetes-informed care.

Semi-structured interviews will be conducted with patients and clinical and non-clinical primary care staff from clusters allocated to the iDiabetes platform and will be carried out in three rounds. An initial round of interviews will be carried out to coincide with the phased implementation of iDiabetes, the second round of interviews will target participants after their first annual review following iDiabetes implementation, and the third round of interviews target participants after their second annual review under iDiabetes care. For patients’ interviews, the focus will be on their experiences of iDiabetes and its impact on them and their diabetes care. The interviews with staff will explore their views and experiences of implementing and using iDiabetes in their clinical practice. Qualitative data collected will form part of a process evaluation of the iDiabetes intervention and insights gained will help inform future refinements to the platform.

Audio-recorded data from the interviews will be transcribed verbatim. Any relevant documentary data (e.g. guidance, policies, training materials) will be included in the qualitative dataset. All qualitative data will be stored in encrypted folders on secure University of Dundee servers.

### Health economics data collection and management

Alongside clinical effectiveness, the iDiabetes platform must also represent value for money to the health care system. To assess this, a cost-effectiveness health economic evaluation will be conducted using the quantitative study date.

If implemented, the iDiabetes platform will affect treatment plans for patients. For any predicted improvement in health outcomes to be achieved, patient adherence to the treatment plan is essential. To gauge uptake of and adherence to the iDiabetes platform, patient preferences will be explored using the discrete choice experiment (DCE) methodology. DCEs have been widely used in health economics to assess patient and public preferences, including preferences for personalised diabetic management.(20) In the developmental stages of the DCE, ‘think aloud’ interviews will be held with a sub-sample of the study population to test and refine the survey. DCE data collection will involve inviting participants to complete an online survey.

No personally identifiable information will be collected in the DCE survey. Data generated by the DCE will be stored on secure University of Aberdeen servers, in password-protected folders which are only accessible to the research team.

### Data Analysis

#### Primary statistical analysis

The primary composite endpoint will be analysed using the Win-Ratio (WR) method. This approach is hierarchical, allowing for sequential analysis of pre-specified endpoints within the composite in order of clinical importance. The win-ratio allows for a combination of quantitative outcomes, such as HbA1C and eGFR reduction, with time-to-first occurrence outcomes such as time to death or hospital admission, in contrast to other conventional statistical methods that only allow time-to-first occurrence of any event in the composite. (21)

In this study the most clinically important endpoint, all-cause mortality, will first be compared between a patient in one study arm to a patient in the comparator arm. If a patient reaches this endpoint first the result will be considered a “loss” for that patient’s arm and a “win” for the comparator arm and no further endpoints are assessed for that pairing. If neither patient “wins” for this outcome, then the next highest priority endpoint will be compared, until all endpoints have been compared sequentially (see *Primary outcome* for endpoint hierarchy). If neither patient in a pairing “wins” according to any of the endpoints, then that pairing is considered a “draw”. The statistical analysis will compare every patient in one arm with every patient in the comparator arm. For example, if comparing *iDiabetes (guideline support)* to *iDiabetesPlus* then all 7500 patients in one arm will be compared to all 7500 patients in the comparator arm, for a total of 56,250,000 opportunities for “wins”, “losses” or “draws”. The total number of “wins” for each arm will be calculated. An arm will be considered superior to the comparator if there are statistically significant increased number of “wins” in that arm compared to the comparator arm. The WR will be estimated and statistical significance will be considered for the primary outcome at the 5% level.

For the statistical analysis, in a Hochberg hierarchical procedure the primary comparison will first be made between the *iDiabetesPlus* and *usual diabetes care* arms. If there is a significant ‘win’ in the *iDiabetesPlus* arm compared to *usual diabetes care* for the composite primary study outcome, comparison will then be made between the *iDiabetes* (guideline support) and *usual diabetes care* arms. If the iDiabetes (guideline support arm) is also demonstrated to be superior to the *usual diabetes care* arm, then a final comparison between the *iDiabetesPlus* and *iDiabetes* (guideline support) arms will be undertaken. This will inform whether enhanced phenotyping and genotyping provide additional advantages over conventional treatment recommendations based on guidelines.

#### Secondary statistical analysis

The main secondary analyses will consist of the comparisons between arms for individual components of the composite, i.e. all-cause mortality, all-cause hospitalisation rate, proportion with >40% eGFR reduction from baseline, new development of ESRD, and proportion with absolute HbA1C reduction >0.5% (>5.5mmol/mol). Secondary outcomes as listed in the study outcomes above will also be analysed treating these as exploratory analyses.

All analyses will be on an intention to treat basis.

A full statistical analysis plan will be published prior to data-lock.

#### Qualitative data analysis

Interview transcripts will be checked for accuracy, anonymised, and then imported into QSR NVivo software to aid analysis and management of data. Thematic analysis will be used to identify and analyse patterns and themes across the dataset.(22) Analysis will take place concurrently with the data collection process to enhance rigour and trustworthiness of findings. Additional techniques to enhance trustworthiness will be put in place, including independent coding triangulation and group-based data analysis critique and data interpretation sessions with the rest of the research team and the study’s PPI group.

#### Health economics analysis

A model-based cost-effectiveness analysis of the *iDiabetes (guideline support)* and *iDiabetesPlus* arms versus *usual diabetes care* arm will be conducted. The cost per complication avoided and Quality-Adjusted Life Year gained (QALYs – the standard health economic utility measure) for each strategy will be predicted by incorporating the study data into an existing diabetes simulation model.(23–25) To estimate the resource implications of introducing the iDiabetes platform at scale in NHS Scotland, a budget impact analysis will be conducted. This will assess whether the recommendations from the cost-effectiveness analysis model are affordable to NHS Scotland.

The DCE data will be used to: (i) assess the relative importance of attributes in the delivery of personalised diabetes care; (ii) predict patient uptake of alternative treatment plans and (iii) estimate benefit-risk trade-offs associated with the risk of adverse outcomes. The DCE data will complement the cost-effectiveness analysis evidence, ensuring that the iDiabetes platform is valued by service users.

#### Ancillary analyses

Exploratory objectives will be evaluated through dedicated sub-studies. The methodology for analysis will be disseminated through the publications of each sub-study.

### Sample size calculations

#### Simulation of power for composite outcome

For the composite primary outcome, no standard method has yet been developed for sample size estimation, but Zhang and Jeong have developed an R-package which simulates the cluster Win-Ratio (cWR) which has been adapted for this study design.(26) The function uses two outcomes - mortality rate and non-fatal hospitalisation. As the primary endpoint in this study is a composite of four outcomes, it will have greater power than that estimated by the simulation as power increases with the number of outcomes in the composite. In the simulations using the R-package developed by Zhang and Jeong, 10% of the planned iDiabetes study population size was used so that there were 60 clusters and 2,400 patients i.e. 40 per cluster and 80 person-years. The gamma frailty parameter beta for shape and rate was set at 20 which is equivalent to an intra-cluster correlation (ICC) = 0.05. The results of 500 simulations gave the proportion of statistically significant results (p<0.05) as 413 so power was 82.6% to detect a mean WR = 1.20. Hence power is excellent using the total number of clusters (60) and only with 10% of the proposed study population size.

#### Power for individual outcomes

In addition, power was also estimated for the individual components of the composite. Difference in Poisson rate or incidence rate ratio was used in these calculations as this is similar to hazard ratio (HR) in a time to event analysis, especially with low censoring and mortality. For example, for hospitalisation with 40 clusters 81% power is achieved to detect a difference in Poisson rate of 8% or more (45% reduced to 37%) assuming an ICC = 0.03 and alpha = 0.05.

All individual outcome sample sizes were estimated using PASS 2023 Sample Size and Power analysis software.

#### Qualitative study sampling

A purposeful, maximum variation sampling strategy will be employed. Initial sampling criteria will account for characteristics relating to setting, including type of intervention (*iDiabetes (guideline support)* or *iDiabetesPlus* settings), practice size (<6000, 6000+) and deciles of deprivation (1–3, 4–7, 8–10); and participants, including staff (e.g. seniority, professional groups, roles) and patients (age, T1D or T2D) to maximise variation. Sampling criteria will be subsequently refined as data collection and data analysis progress. The estimated overall sample size is 20-25 primary care staff and 38-46 patients. Recruitment will take place in primary and secondary care iDiabetes clinical visits, and via recruitment adverts on the iDiabetes and MyDiabetesMyWay websites. If further recruitment avenues are required, SHARE (Scottish Health Research Register and Biobank) will be used to identify and approach potential participants. Healthcare professionals will be invited via professional care networks and internal email lists.

#### Discrete Choice Experiment

As with the qualitative study, a purposeful, maximum variation sampling strategy will be employed. Think aloud interviews will be held until saturation point is reached (when two consecutive interviews yielded no new information regarding survey improvements).(27, 28) It is anticipated saturation will be reached with 20 interviews. Minimum sample size for the DCE survey is calculated using Louviere’s formula for choice proportions. Given a baseline choice probability of 33% (as each choice will involve three options), an accuracy level of 90%, a confidence interval of 95% and eight choice tasks per respondent, we require 234 respondents.(29) We will explore preference heterogeneity using flexible logit models, therefore a minimum of 5000 individuals will be sent an email link to the survey. Assuming a response of 30%, this will allow for sub-group analysis (how preferences differ according to characteristics of respondents). We will recruit at least 234 individuals to pilot the survey.

### Data monitoring

Data monitoring will be undertaken by the study management board. The study has no data monitoring committee as this is considered a low-risk study. This was discussed with the study Scientific Advisory Board who agreed with this approach.

### Adverse event reporting

As all patients’ care is managed by their usual diabetes care teams and the iDiabetes platform only generates treatment recommendations, there will be no adverse event reporting.

## PATIENT AND PUBLIC INVOLVEMENT

A patient and public involvement (PPI) group was formed specifically for this study, consisting of patients and family members recruited from the NHS Scotland Diabetes Research Register and via social media outreach. The PPI group was consulted on all aspects of the project including shaping of the study concept prior to funding application. Subsequently, two representatives were invited from the group as the study’s co-applicants for funding and ethics application and are part of the Study Management Board. Further regular meetings were organised via remote video conference to seek feedback from the PPI group on study design, patient-facing materials including participant information sheets and recommendation outputs which will be viewed by patients on the My Diabetes My Way patient interface as well as study website content. [See Figure 4] Additional patient feedback will be sought as part of the analysis of the platform, described in the qualitative analysis section above.

**Figure 4:**
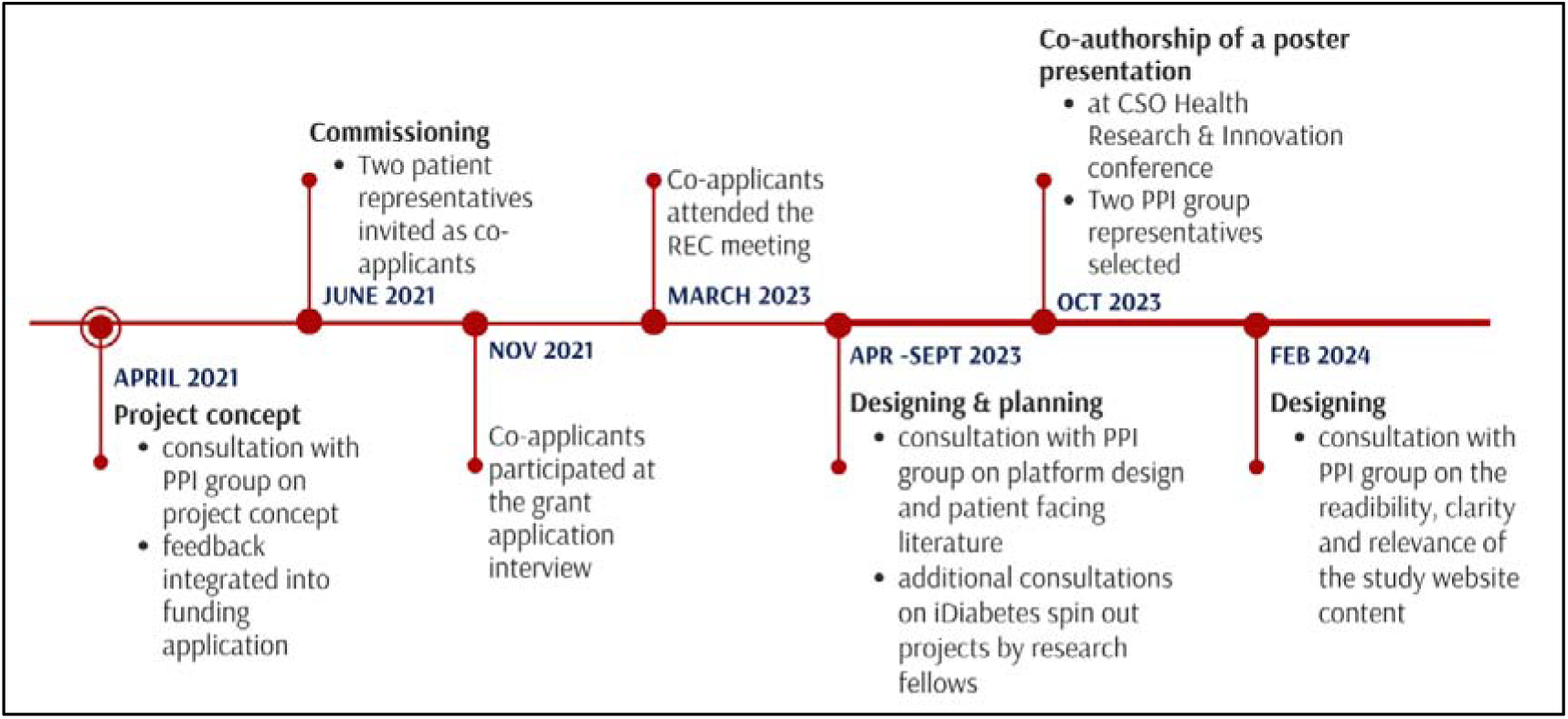
An overview and timeline of the PPI activities conducted throughout the iDiabetes study process.

## ETHICAL CONSIDERATIONS & DISSEMINATION PLAN

### Consent

#### iDiabetes study consent

Consent to take part in the study will take place at a cluster level on randomisation to an iDiabetes platform arm (*iDiabetes (guideline support)* or *iDiabetesPlus)*. Patients will be sent information about the iDiabetes study if their primary care GP is randomised to the *iDiabetes (guideline support)* or *iDiabetesPlus* arm. Patients will not be required to consent individually. Patients who are registered with a primary care GP randomised to an iDiabetes platform arm will be given the opportunity to opt out of the study at an individual level at any point throughout the study period. This can be done when they attend for their annual diabetes review. This approach was deemed acceptable by both the iDiabetes PPI group and the NHS Research Ethics Committee on the grounds that the study intervention was in the form of treatment recommendations, *not* direct changes to treatment and that these recommendations are based on international diabetes guidelines on standard of care.

#### Genetic testing consent

To comply with the requirements of the Human Tissue Act 2004, explicit individual patient verbal consent for genetic testing will be obtained from patients registered with primary care GPs randomised to the *iDiabetesPlus* arm prior to genotyping. This will take the form of the question “*Do you agree to your blood sample undergoing genetic analysis as part of iDiabetes and for future clinical care?*”, with the answer recorded in the SCI-Diabetes system by the person taking the blood test. Patients who do not wish to undergo genetic testing can still take part in the *iDiabetesPlus* arm without this aspect.

#### Echocardiography consent

For iDiabetesPlus patients with a pro-BNP 400 to 2000 pg/ml, further evaluation for heart failure will be performed using AI-assisted echocardiography using US2Ai echocardiography software. An additional PIS will be sent and informed consent obtained for patients prior to undergoing US2Ai echocardiography. For those who do not wish to consent they will be offered a routine NHS echocardiography appointment.

#### Qualitative study consent

Additional informed consent will be taken from participants who are invited to participate in semi-structured interviews as part of the qualitative studies.

#### Health economics study consent

Informed consent will be taken from participants prior to the think aloud interviews and DCE survey.

See Supplemental File for consent documents.

### Research ethics committee approval

This study was reviewed by the NHS Health Research Authority and given a favourable opinion by the East of Scotland Research Ethics Committee (REC reference: 23/ES/0008).

### Confidentiality

The study team will uphold the requirements of legal regulations such as the General Data Protection Regulation (EU) 2016/679 (GDPR), UK Data Protection Act 2018 and NHS Scotland Code of Practice on Protecting Participant Confidentiality when conducting the study. Published results will not contain any personal data that could allow identification of individual participants.

### Dissemination Plan

The criteria for authorship will follow the criteria of the International Committee of Medical Journal Editors. Study findings will be published with the intention to submit a manuscript for publication no later than 12 months after the end of study. Planned dissemination includes the following manuscripts: the main iDiabetes quantitative finding, iDiabetes qualitative analysis, iDiabetes cost-effectiveness analysis and iDiabetes DCE. Additional manuscripts may be prepared on aspects of the statistical analysis, technical development of the iDiabetes platform and ancillary outcomes. Study findings will also be submitted as abstracts to be presented at relevant scientific conferences.

A newsletter giving a summary of the results of the study will be made available to patients on the study website. There will be involvement with local and social media so patients with diabetes in Tayside are aware of the initiative and what it delivers.

### Access to data and amendments

Study-related monitoring, audits, REC review, and regulatory inspection will be permitted. In the event of an audit, the Sponsor, representatives of the Sponsor or regulatory authorities will be given direct access to all study records.

Individual level patient data will not be made publicly available due to data privacy/GDPR regulations. Additional access to the final study dataset on Health Informatics Centre Trusted Research Environment (University of Dundee) will be approved by the chief investigator with an appropriate data sharing agreement in place.

Amendments to the protocol will not be implemented without approval from the Sponsor and subsequent approval from the appropriate REC and NHS R&D Office. Any major amendments undertaken between the publication of the study protocol and the end of the study will be made explicit in the final published study report.

### Post-study care

All diabetes care will be provided by the patients’ diabetes care team throughout the study with recommendations provided by iDiabetes platform. Following the end of the study, no further recommendations will be made by iDiabetes.

### Study Sponsor

University of Dundee, no. 2-026-22. Contact

### Funding

This work was supported by Chief Scientist Office, Scotland (grant number: PMAS-21-01).

The study sponsor & funder was not involved in the study design and will not be involved in the collection, management, analysis, and interpretation of data; writing of the report; and the decision to submit the report for publication nor will they have any have ultimate authority over any of these activities

### Contributors

The study concept and design were conceived by ERP, SB, SGC, TC, JFD, PTD, CL, IRM, HCM, MR, RH and consulted with the PPI group. SC led the development of the iDiabetes IQ engine and dashboard. Quantitative analysis and interpretation will be performed by PD and HW. Qualitative analysis work package components will be coordinated by AF with mixed methods integration performed with TC. Health economics analysis will be performed by MR, RH and MA. SMcK is the programme manager. SM acted as iDiabetes PPI group representatives during funding and ethics application. YYL and DL contributed equally to this manuscript. All authors reviewed the manuscript for intellectual content and approved the final version of the manuscript.

### Collaborators

The iDiabetes study team: B Allardice, N Andrew, E Anyebe, A Barnett, D Baty, S Boyd, L Bremner, P Brennan, J Dickson, A Doney, L Donnelly, E Dow, J Francis, V Godfrey, D Kidd, J McLean, L McNeish, E Middleton, C Palmer, R Petty, S Phillip, E Riches, S Srinivasan, P Sweeten, A Taylor, W Urquhart, A Waheed and K Wilson. The study team acknowledges the collaborative effort of NHS Tayside, My Diabetes My Way, MyWay Digital Health, NHS Education for Scotland (NES), NHS Research Scotland (NRS), SCI-Diabetes, University of Aberdeen and University of Exeter.

### Study management board

This consists of clinicians involved in the development of the study design, collaborators, PPI group representatives and the programme manager.

## Acknowledgements

The study team acknowledges the financial support of NHS Research Scotland (NRS), through Diabetes Network and to the iDiabetes PPI group and representatives for their invaluable contributions to the study. We thank Dr.Anna Barnett (NRS) for her assistance with organising PPI activities. Tayside Clinical Trials Unit (TCTU) is acknowledged for assisting with randomisation of the clusters. The Tayside Local Medical Committee GP sub-group provided valuable input from a primary care perspective. Additionally, we extend our deepest gratitude to Mr. Gerry Tosh (one of the iDiabetes PPI representatives) whose recent passing deeply saddened us. His dedication and insights have been invaluable to the study team and he will be fondly remembered.

## Protocol version

V3.0, 22/09/2023

## Study registration

ISRCTN18000901

## Notes

**Competing interests:** YYL, DL, MA, RB, TC, JFD, PTD, AF, RH, SMcK, SM, MR, GT, HW and MW have no competing interests. ERP has received honoraria for speaking from Novo Nordisk, Lilly and Illumina. SB has received consultancy fees from Astra Zeneca, Bayer and GSK. SGC is a director and employee of MyWay Digital Health. CL has received consultancy fees from Amarin, Aztra Zeneca, Boehringher Ingelheim, Novartis and Vifor; and research grants from Applied Therapeutics, Anacardia, Astra Zeneca, British Heart Foundation, Boehringer Ingelheim, Chief Scientist Office, Eli Lilly, Horizon 2020 EU funding, JDRF, Moderna, NIHR-HTA, Roche Diagnostics, Novo Nordisk, Novartis, and UKRI. HCM has received honoraria from Novo Nordisk and Astra Zeneca. IRM has received honoraria from Astra Zeneca and Boehringer Ingelheim. DJW is a shareholder and cofounder of MyWay Digital Health.

### Competing Interest Statement

YYL, DL, MA, RB, TC, JFD, PTD, AF, RH, SMcK, SM, MR, GT, HW and MW have no competing interests.
ERP has received honoraria for speaking from Novo Nordisk, Lilly and Illumina.
SB has received consultancy fees from Astra Zeneca, Bayer and GSK.
SGC is a director and employee of MyWay Digital Health.
CL has received consultancy fees from Amarin, Aztra Zeneca, Boehringher Ingelheim, Novartis and Vifor; and research grants from Applied Therapeutics, Anacardia, Astra Zeneca, British Heart Foundation, Boehringer Ingelheim, Chief Scientist Office, Eli Lilly, Horizon 2020 EU funding, JDRF, Moderna, NIHR-HTA, Roche Diagnostics, Novo Nordisk, Novartis, and UKRI.
HCM has received honoraria from Novo Nordisk and Astra Zeneca.
IRM has received honoraria from Astra Zeneca and Boehringer Ingelheim.
DJW is a shareholder and cofounder of MyWay Digital Health.

### Clinical Trial

ISRCTN18000901

### Funding Statement

This work was funded by Chief Scientist Office, Scotland (grant number: PMAS-21-01).

### Author Declarations

The East of Scotland Research Ethics Committee (NHS Scotland) gave ethical approval for this work.

### Summary of Updates

Manuscript title: to include study location Abstract 'Methods and Analysis': to include follow-up period Qualitative data collection and management: to clarify the content of the interviews, and the areas that the qualitative data will inform Quantitative data collection and management; Secondary outcomes: data and method used for assessing drug adherence clarified iDiabetes IQ engine performance testing: grammatical error NO CHANGE TO PREVIOUSLY SUBMITTED SUPPLEMENTAL FILES

