## Supplemental File for "The iDiabetes Platform: Enhanced Phenotyping of Patients with Diabetes for Precision Diagnosis, Prognosis and Treatment - study protocol for a cluster-randomised controlled study in Tayside, Scotland" (from another version of this preprint: version 1, 2024-03-21)

**Exploratory objectives and outcomes**

*Exploratory objectives*

- Assess the impact of implementing a precision diabetes care model on the incidence and progression of renal disease
- Assess the impact of implementing a precision diabetes care model on Major Adverse Cardiac Outcomes (MACE)
- Assess the impact of implementing a precision diabetes care model on the early detection of subclinical cardiovascular disease and all phenotypes of heart failure
- Assess the impact of implementing a precision diabetes care model on the diagnostic frequency of metabolic dysfunction-associated steatotic liver disease (MASLD) with advanced fibrosis or cirrhosis
- Understand the organisational context and practices involved in the implementation and delivery of iDiabetes platform
- Explore the impact of implementing iDiabetes platform on primary and secondary care provision from the perspective of patients and health professionals
- Evaluate the cost-effectiveness, budget impact, and patient preferences for the precision medicine platform
- Assess the impact of socio-economic deprivation on outcomes of a precision diabetes care model

*Exploratory outcomes (measured at median of 2 years from the start of recruitment unless specified otherwise)*

1. Number of patients with onset of acute kidney injury (AKI) – defined as an increase in serum creatinine ≥ 1.5 times the latest value (known or presumed to have occurred within the prior 7 days) (as defined by 2012 KDIGO guideline) (1)
2. Number of patients initiating on kidney replacement therapy due to AKI
3. Number of patients with albumin:creatinine ratio (ACR) >20mg/mmol
4. Number of patients with albumin:creatinine ratio (ACR) >3mg/mmol
5. Mean annual rate of change in eGFR from baseline to final follow-up measure (in those with eGFR<60ml/min at baseline)
6. MACE outcomes stratified by drug therapy
7. New diagnoses of heart failure (with preserved and reduced ejection fraction)
8. Change in cardiovascular risk (UKPDS 10-year CV risk)
9. Number of patients with ALT >30 IU/L
10. New diagnoses of MASLD with or without advanced fibrosis or cirrhosis
11. New diagnoses of liver disease secondary to other aetiologies
12. Progression/regression of non-invasive fibrosis scores and liver stiffness (based on Fibroscan finding) stratified by drug therapy
13. HbA1C level at initiation of drug therapy (treatment inertia)
14. Change in HbA1C at end of study
15. Change in HbA1C at 6 months after initiation of new drug therapy
16. Antidiabetic medications cessation rate due to insufficient glycaemic response
17. Adherence of new drug therapy for more than 6 months
18. Number of patients with change in diabetes diagnosis
19. Number of patients with new onset diabetes-related eye disease (retinopathy or maculopathy)
20. Hospitalisation rate secondary to diabetes-related foot disease

21. Amputation rate secondary to diabetes-related foot disease
22. Acceptability of iDiabetes platform (from the perspective of different users) and their adaptability to the intervention
23. Cost per diabetes-related complication avoided
24. Quality-Adjusted Life Year gained (QALYs) for each iDiabetes intervention arm
25. Utilising discrete choice experiments: (i) assess the relative importance of attributes in the delivery of personalised diabetes care; (ii) predict patient uptake of alternative treatment plans and (iii) estimate benefit-risk trade-offs associated with the risk of adverse outcomes
26. Impact of socio-economic deprivation (based upon SIMD) on the following outcomes: primary composite endpoint, hospitalisation rate, proportion with >40% eGFR reduction from baseline, or ESKD, drug adherence rate, change in HbA1C upon new drug treatment initiation and at end of study
