## Supplemental File for consent documents for "The iDiabetes Platform: Enhanced Phenotyping of Patients with Diabetes for Precision Diagnosis, Prognosis and Treatment - study protocol for a cluster-randomised controlled study in Tayside, Scotland" (from another version of this preprint: version 1, 2024-03-21)

### The iDiabetes Platform: the views and experiences of health professionals and patients

Consent form: Patient interviews Participant ID_________
Chief Investigator: Prof Ewan Pearson, Professor of Diabetic Medicine
Sponsor: University of Dundee

Please initial box for each statement below:

| Statement | Initials |
|---|---|
| 1. I have read and understood the information sheet [version__ /date__ ] for the research project entitled "The iDiabetes Platform: the views and experiences of health professionals and patients". | |
| 2. I have had enough time to consider the information and to ask questions, and I am happy with any answers I have been given. | |
| 3. I understand that my participation in the interview(s) is voluntary and that I am free to withdraw my participation at any time, without giving any reasons. I know this will not affect my care or legal rights. | |
| 4. I agree to the researcher audio-recording the session. | |
| 5. I understand that data collected during the study will be looked at by the research team, and that all information will remain anonymous and confidential, and that no personal information will be used which may identify me or any other people in the final report or scientific publications. | |
| 6. I understand that all data collected (e.g. audio-recordings) will be stored in a secure University server and line with current Data Protection regulations. | |
| 7. I understand that data collected during the study may be looked at by individuals from regulatory authorities who have a duty to monitor the quality of the research, and I give permission for this. | |
| 8. I understand that if any disclosures are made during the sessions that suggest malpractice, misconduct, or that someone is in danger of harm, this information will be shared with the appropriate personnel. | |
| 9. I agree to take part in this research study. | |

Name of participant (capitals) Date Signature

_______________________ _________ _______________

Name of person taking consent (capitals) Date Signature

_______________________ _________ _______________

### The iDiabetes Platform: the views and experiences of health professionals and patients

Consent form: Staff interviews — Participant ID_________

Chief Investigator: Prof Ewan Pearson, Professor of Diabetic Medicine

Sponsor: University of Dundee

Please initial box for each statement below:

| Statement | Initials |
|---|---|
| 1. I have read and understood the information sheet [version/date] for the research project entitled "The iDiabetes Platform: the views and experiences of health professionals and patients". | |
| 2. I have had enough time to consider the information and to ask questions, and I am happy with any answers I have been given. | |
| 3. I understand that my participation in the interview(s) is voluntary and that I am free to withdraw my participation at any time, without giving any reasons. I know this will not affect my care or legal rights. | |
| 4. I agree to the researcher audio-recording the session. | |
| 5. I understand that data collected during the study will be looked at by the research team, and that all information will remain anonymous and confidential, and that no personal information will be used which may identify me or any other people in the final report or scientific publications. | |
| 6. I understand that all data collected (e.g. audio-recordings) will be stored in a secure University server and line with current Data Protection regulations. | |
| 7. I understand that data collected during the study may be looked at by individuals from regulatory authorities who have a duty to monitor the quality of the research, and I give permission for this. | |
| 8. I understand that if any disclosures are made during the sessions that suggest malpractice, misconduct, or that someone is in danger of harm, this information will be shared with the appropriate personnel. | |
| 9. I agree to take part in this research study. | |

| Name of participant (capitals) | Date | Signature |
|---|---|---|
| ______________________ | __________ | ______________________ |

| Name of person taking consent (capitals) | Date | Signature |
|---|---|---|
| ______________________ | __________ | ______________________ |

### iDiabetes: Echo scan

Consent form:  iDiabetes: Echo Scan  Participant ID__________
Chief Investigator: Prof Ewan Pearson, Professor of Diabetic Medicine
Sponsor: University of Dundee

Please initial box for each statement below:

| Statement | Initials |
|---|---|
| 1. I have read and understood the participant information sheet [version__ /date__ ] for the research project entitled "iDiabetes Study: Echo Scan". | |
| 2. I have had enough time to consider the information and to ask questions, and I am happy with any answers I have been given. | |
| 3. I understand that my participation in the study is voluntary and that I am free to withdraw my participation at any time, without giving any reasons. I know this will not affect my care or legal rights. | |
| 4. I agree to have an echo scan using US2Ai software. | |
| 5. I understand that data collected during the study will be looked at by the research team and that all information will remain confidential, and that no personal information will be used which may identify me or any other people in the final report or scientific publications. | |
| 6. I understand that the echo results will be shared with my clinical care team including my GP where appropriate. | |
| 7. I understand that data collected will be stored in secure University servers in line with current Data Protection regulations. | |
| 8. I understand that data collected during the study may be looked at by individuals from regulatory authorities who have a duty to monitor the quality of the research, and I give permission for this. | |
| 9. I agree to take part in this research study. | |

| Name of participant (capitals) | Date | Signature |
|---|---|---|
| ______________________ | __________ | ______________________ |
| Name of person taking consent (capitals) | Date | Signature |
| ______________________ | __________ | ______________________ |
